# Lactoferrin and lysozyme for Kenyan children presenting with wasting and diarrhea: A 2 x 2 factorial randomized controlled trial

**DOI:** 10.64898/2026.04.27.26351844

**Authors:** K.D. Tickell, R. Tiwari, I. Trehan, J. Otieno, M. Okello, A. Shah, L. Keter, E. Yoshioka, E. Ochola, C. Nyabinda, D. Rwigi, J.M. Njunge, E. Houpt, J.A. Platts-Mills, J. Liu, B.A. Richardson, C.J. McGrath, AR Means, M.M. Diakhate, G. John Stewart, J.L. Walson, B.O. Singa, P.B. Pavlinac

## Abstract

**Introduction:** Lactoferrin and lysozyme are milk derived proteins with antimicrobial and anti-inflammatory properties. We tested if these supplements improved time-to-nutritional-recovery and reduced the incidence of new moderate or severe diarrhea (MSD) among children presenting to hospital with wasting and diarrhea.

**Methods:** Medically-stable children aged 6-24 months with diarrhea and wasting were randomized to a 16-week course of lactoferrin, lysozyme, a combination of both, or placebo. Time-to-nutritional-recovery (mid-upper arm circumference ≥ 12.5cm) and incidence of new onset MSD were the primary outcomes observed over 6-months follow-up. Subgroup analyses included efficacy by wasting status (severe vs. moderate), stunting, age, inpatient/outpatient, and adherence.

**Results:** Among the 600 children randomized, 531 (88.5%) nutritionally recovered within 16-weeks; 63% among severely wasted children and 95% in children with moderate wasting. The wasting recovery rate in the combination arm was non significantly higher (HR: 1.23, 95%CI: 0.97, 1.57; p=0.083) than the placebo group. Children randomized to lactoferrin alone and lysozyme alone had nutritional recovery rates similar to placebo (HR: 0.94, 95% CI 0.74, 1.20; p=0.607 and HR: 0.91, 95% CI: 0.71, 1.17; p=0.462, respectively). Among severely wasted children, the combination arm had a higher recovery rate than placebo (HR: 2.76, 95% CI 1.49, 5.09; p=0.001), but not the individual lactoferrin (HR: 1.29, 95% CI 0.69, 2.41; p=0.427) and lysozyme (HR: 0.80, 95% CI: 0.40, 1.60; p=0.530) arms. Children randomized to intervention arms had comparable incidence of MSD (82.3-97.0 per 100 child-years) to the placebo arm (75.3 per 100 child-years).

**Conclusions:** The combination of lactoferrin and lysozyme for 16 weeks modestly improved nutritional recovery time particularly among severely wasted children. If confirmed, there may be a role for enteric-targeted therapeutics as adjuvants to severe wasting management. Additional strategies are needed for the post-acute diarrhea recovery period.

## Introduction

Wasting affects 43 million children each year and children with wasting have a higher incidence of diarrhea and are more likely to die during diarrheal episodes than better nourished peers.^1,2^ Comorbid wasting and diarrhea are important contributors to childhood mortality and morbidity in low- and middle-income countries.^3–5^ Approximately 320,000 children under 5 years due each year from diarrhea, and up to 60% of this mortality is among children with comorbid wasting.^6,7^ In addition to proximal morbidity and mortality attributed to comorbid diarrhea and wasting, both conditions are associated with longer term sequalae including repeated diarrheal episodes, chronic enteric inflammation, and delayed growth recovery. Novel interventions that expedite recovery from wasting and prevent repeated episodes of diarrhea could substantially improve child health.

Lactoferrin and lysozyme are breastmilk proteins with known antimicrobial properties and favorable safety profiles. Lactoferrin sequesters iron in the gut limiting rapid bacterial growth, while lysozyme hydrolyzes bacterial cell wall peptidoglycan. Both enzymes have demonstrated broad antimicrobial effects in in-vitro and animal model studies.^8–12^ In clinically healthy Malawian children, a 16-week course of lactoferrin and lysozyme improved enteric barrier dysfunction,^13^ a key mechanism underpinning poor long-term outcomes of wasting and diarrhea, and decreased the incidence of MAM and hospitalizations.^14^ In two different studies from Peru and China, lactoferrin reduced the cumulative prevalence of diarrhea episodes, including severe episodes in healthy children.^15,16^ However, only a single trial in Peru has tested these products as a therapeutic intervention for diarrhea, and reported a two-day faster recovery from diarrhea with the use of lactoferrin and lysozyme-supplemented oral rehydration solution.^17^ These studies suggest lactoferrin and lysozyme may be viable and safe methods to improve the outcomes of children with wasting and diarrhea.

This blinded, randomized, placebo-controlled trial tested whether lactoferrin and/or lysozyme decrease the time to recovery from wasting and reduces the incidence of secondary diarrhea among Kenyan children who presented for care with moderate or severe wasting and diarrhea.

## Methods

This was a two-by-two factorial, blinded, placebo-controlled, randomized trial to determine the efficacy of lactoferrin and lysozyme supplementation in promoting nutritional recovery and decreasing the incidence of diarrhea among children seen at outpatient clinics or being discharged from hospital following management for comorbid moderate or severe diarrhea and wasting.^18^ Kenyan children aged 6-24 months with a mid-upper arm circumference (MUAC) <12.5cm returning home from an outpatient visit or inpatient stay for diarrhea were randomized to 16-weeks of lactoferrin, lysozyme, a combination of the two, or placebo. Children were recruited from Homa Bay County Teaching and Referral, Kisii County Teaching and Referral, Migori County Referral, Rongo Subcounty, and Isebania Subcounty hospitals all located in the Western part of Kenya, between March 2023 and February 2025.

Children were screened for eligibility on inpatient or outpatient treatment units. Eligible participants underwent informed consent in their preferred language (English, Kiswahili, Kisii, Kuria, or Luo). Consenting eligible families were interviewed to assess the child’s demographic information, medical history (including antibiotic use and any other medical management received in the hospital), and contact information. Clinical and laboratory information was abstracted from medical records and all enrolled participants underwent a physical examination performed by the study clinicians trained in anthropometry. Length and weight were measured by two staff, with a third measurement if there was >0.5 cm (length) or 0.1 kg (weight) difference between measurements. After enrollment, standard of care was followed for all children, which included provision of ready-to-use-therapeutic foods (RUTF) and/or ready-to-use-supplemental foods (RUSF) in keeping with the Kenyan National Integrated Management of Malnutrition Guidelines.^19^ In addition all children received oral rehydration, therapeutic zinc unless on RUTF, and antibiotics for dysentery or suspected cholera as outlined by Kenyan National Diarrhea guidelines.^20^

Block randomization (1:1:1:1) in random sized blocks of no more than 12 was used to assign treatment groups at study enrollment. Treatment allocation was concealed and once assigned, the child’s allocation remained blinded to the participants, study staff, hospital clinicians, and the investigators during data collection and analysis phases of the study. To account for the differences in severity of wasting and dehydration between hospitalized children and those seen as outpatients, we stratified randomization by site of recruitment (i.e. inpatient vs outpatient).

Children received a 16-week course of either lactoferrin, lysozyme, combination therapy or placebo. The interventional products were provided in powder form in plain sachets labeled only with the patient ID, expiration date, administration instructions, and study contact information for cases of suspected adverse events. Fourteen sachets were supplied at enrollment and replenished during home and clinic visits every two-weeks. Pictorial adherence log and daily diarrhea diary cards were completed by caregivers and collected by study staff during follow-up visits, at which times new cards were provided. Caregivers were instructed to use one sachet (containing 41.5g of the product) daily, and to mix the sachet of the investigational product (IP) with 125 mL of porridge provided by the study. Caregivers were instructed to prepare the porridge and cool to an edible temperature, then to mix in the IP. Lactoferrin was given at 1.5 grams per day, and lysozyme was given at 40 g of 0.5% lysozyme per day. These doses were based on efficacy in previous trials of a similar age range^13^. Study community health workers (CHWs) visited the homes of all children daily in the first 2 days of study participation to assist the family in preparing and administering the product. If the child was not hungry during the visit, the mixture of IP and porridge was stored for up to 8 hours until the participant was ready to take it. Adherence was assessed for all study participants with each box (containing 14 sachets) of IP assessed based on the number of returned empty sachets during the home and clinic visits as well as by evaluating the caregiver-reported daily administration from the pictorial adherence logs.

Following the daily home visits for the first two days, the study CHWs conducted further home follow-up at weeks 2, 6, 8, 12, 14, 18, 20, and 22. Enrolled children also returned to the hospital for review by the study clinical team at weeks 4, 10, 16, and 24 post enrollment (with transportation cost reimbursed) in addition to their usual fortnightly nutritional clinic visits (as per standard of care). Caregivers also received daily diarrhea diaries at enrollment after counseling on their use, and a standardized questionnaire was administered at all visits ascertaining health history information including presence/absence of diarrhea and diarrhea severity indicators informed by the diarrhea diary and 14-day caregiver recall. Other information, such as concomitant medication use (including antibiotics) and any other non-scheduled clinic visits or hospitalizations was ascertained during follow-up visits. During scheduled facility-based follow-up visits, clinicians performed a physical exam and obtained anthropometric measurements. Adverse event monitoring included active ascertainment of diarrhea, vomiting, skin rash, lip swelling, difficulty breathing/wheeze, seizure, hospitalization, and/or death and passive ascertainment (open-ended question) of any other potential unanticipated events during all visits. If CHWs suspected the child was having an adverse event during a home visit, they contacted the study physician, and when necessary the child was referred to the study health facility for further review.

### Statistical analysis

The co-primary outcomes were time to nutritional recovery and the incidence of moderate or severe diarrhea (MSD), as outlined in the statistical analysis plan on clinicaltrials.gov NCT05519254. Time to nutritional recovery was defined as the number of days between enrollment and the date of the second of two consecutive MUAC measurements ≥12.5 cm. Participants who did not reach nutritional recovery, due to death, loss to follow-up, or completion of the study prior to reaching recovery were censored at the date of their last follow-up visit. We focused on MUAC instead of weight-for-height z-score because MUAC is less sensitive than weight to hydration status (a common consequence of diarrhea).^21^ The incidence of MSD was defined as the total number of new diarrhea episodes (>48 hours after a diarrhea-free period) deemed moderate or severe, divided by the child-time at risk during the 6-month follow-up period. Time at risk was censored at the date of last follow-up for children who died or were lost to follow-up. MSD was defined using the CODA (Community DiarrhoeA) diarrhea severity score of ≥3 or dysentery (evidence or reported visible blood in stool).^22^ A child was considered lost to follow-up if they did not attend at least one of the follow-up clinic visits (at weeks 4, 10, 16, 24) and were not available at any of the home visits at week 2 or beyond (at 2, 6, 8, 12, 14, 18, 20, and 22 weeks). For the nutritional recovery outcome, participants who were lost-to-follow-up were retained in the intention-to-treat (ITT) analysis, censored at the date of death or withdrawal, and classified as not recovered. For the MSD diarrhea outcome, participants who were lost-to-follow-up were classified as having diarrhea, except for one participant who reported no diarrhea in the daily diarrhea diary prior to death. Person-time for diarrhea analyses was accrued up to just before death or withdrawal.

The primary analysis compared each intervention arm to placebo in an ITT analysis. Rate ratios (RRs) of each intervention arm compared to placebo were estimated using a Poisson model with number of episodes as the outcome and time at risk (defined above) as the model offset. Wald chi-square tests of the two-way intervention arm comparisons were used for hypothesis testing. The median time to nutritional recovery was compared between intervention arms using Kaplan Meier (K-M) survival analysis and associated log-rank tests for each two-way comparison of intervention group to placebo. Cox-proportional hazards regression was used for analysis of time to nutritional recovery with adjustment for inpatient/outpatient status (stratification factor). *A-priori* defined sub-group analyses were conducted to evaluate the effect of interventions in sub-groups a) severe/moderate wasting, b) high/medium/low adherence to the intervention (<85%, 85-99%, >99%), and c) age 6-11 months/12-24 months. To better understand the results, ad-hoc subgroup analyses for stunting vs not stunting and inpatient vs outpatient recruitment were added. Additionally, effect modification by lactoferrin and/or lysozyme was tested using a Wald test. Where there was no evidence of effect modification using a conservative p-value of 0.1, the two lactoferrin and two lysozyme arms were tested independently compared to the non-lactoferrin and non-lysozyme arms, respectively, to determine the efficacy of each supplement. Where effect modification was present each intervention arm was compared to placebo independently. Per protocol analyses were specified *a priori* and excluded children who were ineligible but had been recruited, those who were lost to follow-up (defined as missing all follow-up visits), those who had ≤99% adherence, and those who withdrew consent.

All analyses were performed in R (R version 4.5.2). All caregivers provided full, written informed consent for inclusion in the trial which was approved by the University of Washington Institutional Review Board (STUDY00011759) and Kenya Medical Research Institute’s Scientific and Ethical Review Unit (SERU 4274).

## Results

A total of 1527 children were screened for eligibility (**Figure 1**) and 600 were enrolled, and four participants did not attend any follow-up visits due to death (N = 3) or withdrawal (N = 1). The most common reasons for exclusion were MUAC ≥12.5 cm and being outside the 6-24 month recruitment age range. Baseline characteristics appear balanced across the four randomization arms, each with approximately 150 children (lactoferrin n=149; lysozyme n=149; combination n=152; placebo n=150) (**Table 1).** Over half the children (52%) were aged 6-11 months, 54% were female, and 71% were being partially breastfed at the time of enrollment. One in five (20%) of enrolled children were severely wasted, while 52% were stunted, and the majority lived in households that were moderately or severely food insecure (73%). Children had high adherence to the investigational products over the 16 weeks, with 78% reporting >99% consumption of the total 112 daily sachets while 18% reported consuming 85-99% of the sachets (**Supp Table 1**) with consistent adherence across the four randomization arms. Adherence by empty sachet return was similarly high: 73% returned >99% of the empty sachets while 16% returned between 85-99% empty sachets which was also consistent across arms.

**Figure 1.**
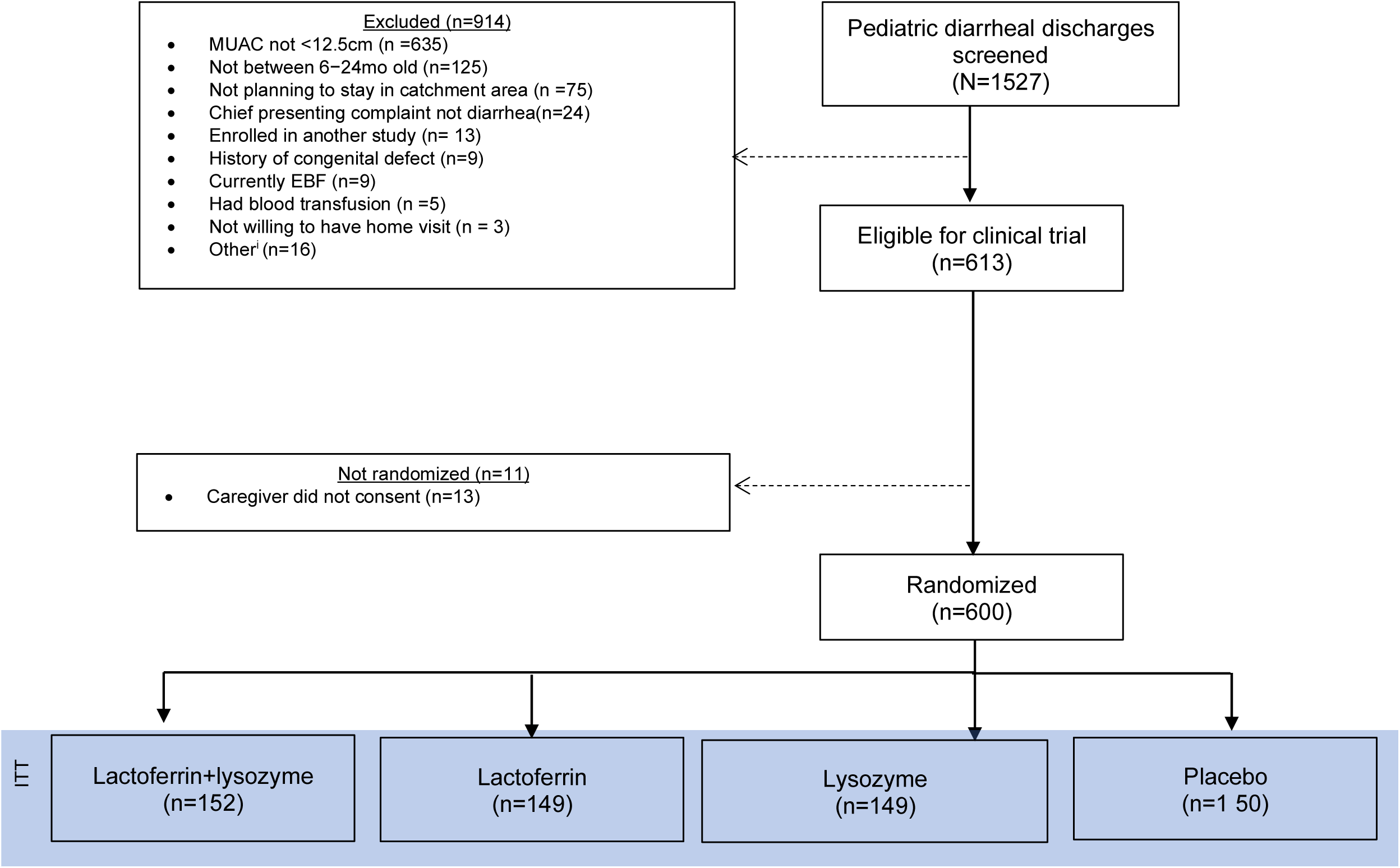
Flowchart of included subjects. ^i^Includes the following exclusions: Child not deemed clinically stable (n =1); Discharged against medical advice (n = 2); Not willing to prepare study product (n=2); Already enrolled in this study (n= 1); Not willing to provide consent (n =2); Not willing to bring child to clinic (n =2); child was already discharged for at least 2 days when screened (n=2); caregiver did not have time to sit through the whole process of consenting (n=1), caregiver was committed elsewhere (n=2), and caregiver requested to consult with partner who declined enrolment (n=1).

**Table 1.** Baseline characteristics by randomization arm.

| Characteristic | Overall (N=600) | Lactoferrin+<br>Lysozyme Arm<br>(n=152) | Lactoferrin Arm<br>(n=149) | Lysozyme Arm<br>(n=149) | Placebo Arm<br>(n=150) |
| --- | --- | --- | --- | --- | --- |
|  | n (%)<br>Median (IQR) | n (%)<br>Median (IQR) | n (%)<br>Median (IQR) | n (%)<br>Median (IQR) | n (%)<br>Median (IQR) |
| Recruitment Site |  |  |  |  |  |
| Kisii Teaching and Referral | 157 (26%) | 41 (27%) | 44 (30%) | 38 (26%) | 34 (23%) |
| Homa Bay County Referral | 176 (29%) | 44 (29%) | 45 (30%) | 41 (28%) | 46 (31%) |
| Rongo Sub-County | 178 (30%) | 45 (30%) | 40 (27%) | 49 (33%) | 44 (29%) |
| Isebania Sub-County | 89 (15%) | 22 (14%) | 20 (13%) | 21 (14%) | 26 (17%) |
| Enrolled from Outpatient Unit<sup>a</sup> | 482 (80%) | 121 (80%) | 120 (81%) | 120 (81%) | 121 (81%) |
| **CHILD** |  |  |  |  |  |
| Age |  |  |  |  |  |
| 6m to 11m | 311 (52%) | 81 (53%) | 71 (48%) | 84 (56%) | 75 (50%) |
| 12m to 17m | 199 (33%) | 51 (34%) | 48 (33%) | 49 (33%) | 51 (34%) |
| 18m to 24m | 90 (15%) | 20 (13%) | 30 (20%) | 16 (11%) | 24 (16%) |
| Median age (months) | 11.6 (9.0, 15.6) | 11.2 (8.8, 15.5) | 12.2 (9.5, 16.4) | 10.9 (8.6, 14.6) | 11.9 (9.2, 15.8) |
| Female | 321 (54%) | 84 (55%) | 75 (50%) | 86 (58%) | 76 (51%) |
| Breastfeeding Status |  |  |  |  |  |
| Ever breastfed | 596 (99%) | 151 (99%) | 148 (99%) | 148 (99%) | 149 (99%) |
| Currently breastfeeding | 426 (71%) | 114 (75%) | 97 (65%) | 118 (79%) | 97 (65%) |
| Exclusively breastfed for 6m<sup>b</sup> | 322 (55%) | 85 (56%) | 80 (54%) | 77 (53%) | 80 (55%) |
| Nutritional Status |  |  |  |  |  |
| Stunted (HAZ < -2) | 314 (52%) | 83 (55%) | 82 (55%) | 72 (48%) | 77 (51%) |
| Severely wasted (MUAC < 12.5cm) | 122 (20%) | 27 (18%) | 34 (23%) | 27 (18%) | 34 (23%) |
| Underweight (WAZ < -2)<sup>c</sup> | 387 (66%) | 97 (65%) | 93 (64%) | 94 (65%) | 103 (71%) |
| MUAC (cm) | 12.20<br>(11.70, 12.30) | 12.20<br>(11.75, 12.30) | 12.20<br>(11.60, 12.30) | 12.20<br>(11.80, 12.30) | 12.20<br>(11.80, 12.30) |
| HIV Status |  |  |  |  |  |
| HIV-infected | 5 (0.8%) | 3 (2.0%) | 1 (0.7%) | 1 (0.7%) | 0 (0%) |
| HIV-exposed uninfected | 26 (4.3%) | 9 (5.9%) | 4 (2.7%) | 5 (3.4%) | 8 (5.3%) |
| HIV-exposed status unknown | 50 (8.3%) | 15 (9.9%) | 9 (6.0%) | 13 (8.7%) | 13 (8.7%) |
| HIV-unexposed | 376 (63%) | 89 (59%) | 95 (64%) | 103 (69%) | 89 (59%) |
| HIV status unknown | 143 (24%) | 36 (24%) | 40 (27%) | 27 (18%) | 40 (27%) |
| **CAREGIVER** |  |  |  |  |  |
| Education (primary or never attended) | 264 (44%) | 63 (41%) | 69 (46%) | 68 (46%) | 64 (43%) |
| Married<sup>d</sup> | 439 (73%) | 119 (78%) | 110 (74%) | 113 (76%) | 97 (65%) |
| Nutritional Status |  |  |  |  |  |
| MUAC (cm) | 26.0 (24.2, 28.6) | 26.1 (24.4, 28.6) | 26.6 (24.5, 29.0) | 25.8 (24.0, 28.3) | 26.0 (24.1, 28.6) |
| Underweight<sup>e</sup> | 251 (42%) | 62 (41%) | 54 (36%) | 69 (46%) | 66 (44%) |
| Normal weight<sup>f</sup> | 231 (39%) | 62 (41%) | 65 (44%) | 52 (35%) | 52 (35%) |
| Overweight<sup>g</sup> | 118 (20%) | 28 (18%) | 30 (20%) | 28 (19%) | 32 (21%) |
| HIV Status |  |  |  |  |  |
| HIV-infected | 81 (14%) | 27 (18%) | 14 (9.4%) | 19 (13%) | 21 (14%) |
| HIV- uninfected | 365 (61%) | 88 (58%) | 92 (62%) | 98 (66%) | 87 (58%) |
| HIV-exposure status unknown | 154 (26%) | 37 (24%) | 43 (29%) | 32 (21%) | 42 (28%) |
| **HOUSEHOLD** |  |  |  |  |  |
| Crowding (≥3 people per room living in house) | 398 (66%) | 104 (68%) | 100 (67%) | 97 (65%) | 97 (65%) |
| Monthly household income<sup>h</sup> <5,000 Kenya Shillings | 320 (53%) | 81 (53%) | 84 (57%) | 77 (52%) | 78 (52%) |
| Toilet type |  |  |  |  |  |
| Flush<sup>i</sup> | 47 (7.8%) | 13 (8.6%) | 8 (5.4%) | 9 (6.0%) | 17 (11%) |
| Pit latrine<sup>j</sup> | 535 (89%) | 135 (89%) | 139 (93%) | 132 (89%) | 129 (86%) |
| Open defecation<sup>k</sup> | 18 (3.0%) | 4 (2.6%) | 2 (1.3%) | 8 (5.4%) | 4 (2.7%) |
| Food insecurity<sup>l</sup> |  |  |  |  |  |
| Secure | 94 (16%) | 27 (18%) | 22 (15%) | 21 (14%) | 24 (16%) |
| Mild insecurity | 66 (11%) | 13 (8.6%) | 18 (12%) | 15 (10%) | 20 (13%) |
| Moderate insecurity | 234 (39%) | 64 (42%) | 56 (38%) | 59 (40%) | 55 (37%) |
| Severe insecurity | 206 (34%) | 48 (32%) | 53 (36%) | 54 (36%) | 51 (34%) |

<sup>a</sup>Includes children in short-term holding rooms or diarrhea treatment clinic
<sup>b</sup>Only among those who have ever breastfed and whose age they were introduced other foods and or liquids is known
<sup>c</sup>Participants who have edema do not have WAZ score calculated.
<sup>d</sup>Includes caregivers who reported being in monogamous or polygamous marriage.
<sup>e</sup>Maternal underweight defined as MUAC ≤ 25.5cm
<sup>f</sup>Maternal normal nutritional status defined as MUAC > 25.5cm and < 29.5cm.
<sup>g</sup>Maternal overweight defined as MUAC ≥ 29.5cm
<sup>h</sup>Defined as participant's total household income in the last month.
<sup>i</sup>Defined as flushed or flush that are: pour to piped sewer system, pour to septic tank, pour to somewhere else, or don't know where.
<sup>j</sup>Defined as pit latrines: ventilated improved, with slab floor, or without slab floor.
<sup>k</sup>Defined as open defecation/bush/field
<sup>l</sup>Defined using the 'Household Food Insecurity Access Scale (HFIAS) for Measurement of Food Access' (v3, 2007)
Abbreviations: HAZ: length/height-for-age z-score; MUAC: Mid-upper arm circumference; WAZ: weight-for-age z-score.

### Recovery from wasting

Of the 600 children included in the ITT analysis, 531 (89%) nutritionally recovered within 16-weeks (63% among severely wasted children and 95% in children with moderate wasting, **Figure 2, Supp Table 2**). The primary wasting outcome, recovery rate, in the placebo arm was 489.4 per 100-child years, while the combination arm had a modestly higher recovery rate (599.9 per 100 child-years; HR: 1.23, 95%CI: 0.97, 1.57; p=0.083, **Table 2**). Children randomized to either lactoferrin or lysozyme had similar nutritional recovery rates to those in placebo (HR: 0.94, 95% CI 0.74, 1.20; p=0.607 and HR: 0.91, 95% CI: 0.71, 1.76; p=0.462, respectively). Because there was evidence of synergy between the two supplements (Wald test) for the interaction term p=0.036), we did not combine the lactoferrin or lysozyme arms.

**Figure 2:**
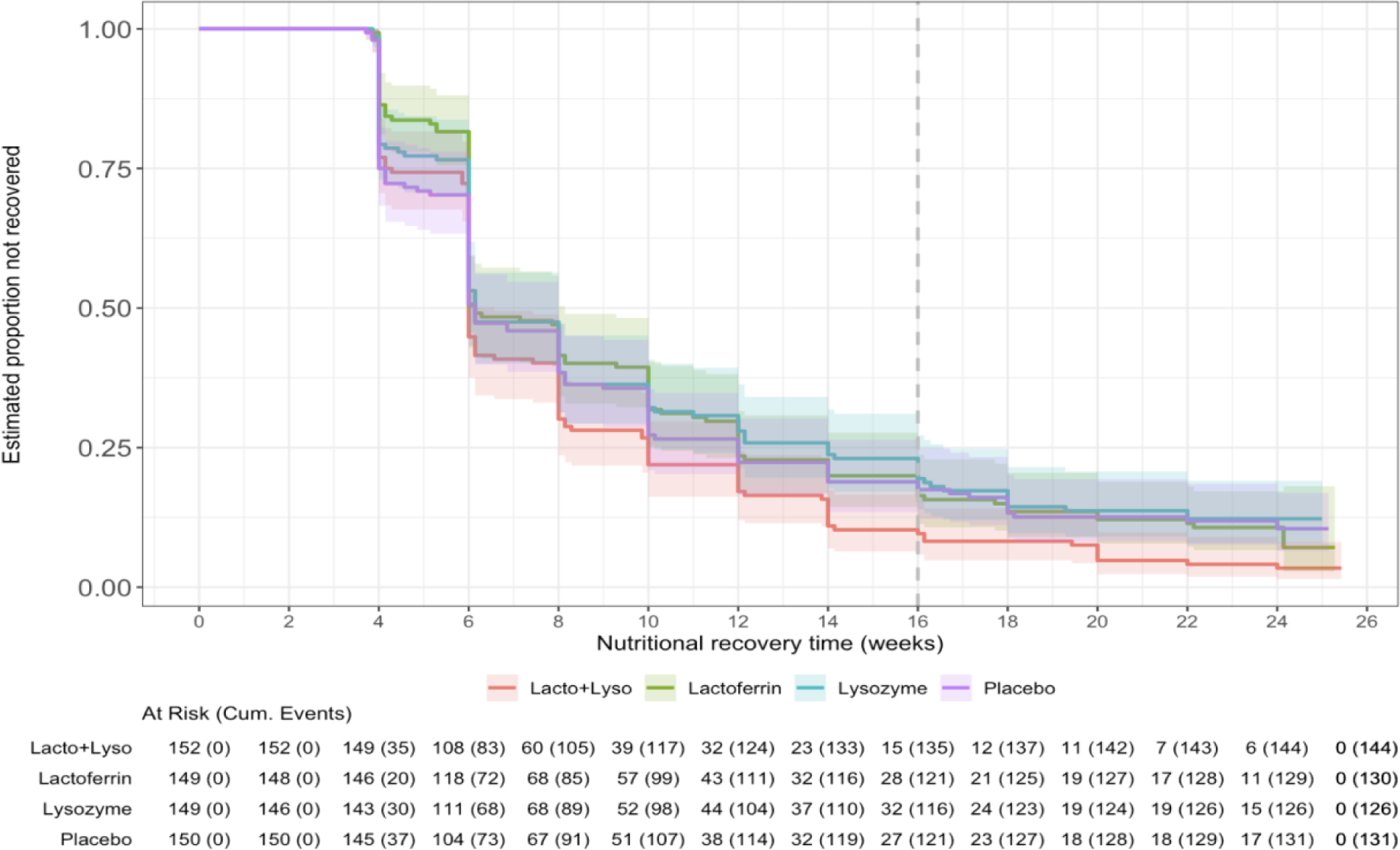
Kaplan-Meier curve for time to nutritional recovery, by randomization arm. Kaplan–Meier curves are shown for the number of days until nutritional recovery was achieved or until the last follow-up visit occurred (for those who did not recover during the study and if follow-up visit <183 days). Nutritional recovery defined as the number of days since enrollment to the date of the 2nd of two consecutive fortnightly MUAC measurements ≥12.5 cm. Dashed line at week 16 denotes end of the nutritional intervention.

**Table 2.** Primary and secondary outcomes Nutritional recovery and moderate-to-severe diarrhea and by randomization arm.

| Outcome | Lactoferrin+Lysozyme Arm |  |  |  |  | Lactoferrin |  |  |  |  | Lysozyme Arm |  |  |  |  | Placebo Arm |  |  |
| --- | --- | --- | --- | --- | --- | --- | --- | --- | --- | --- | --- | --- | --- | --- | --- | --- | --- | --- |
|  | n<sup>i</sup> | Person-time<sup>ii</sup> | Incidence rate<sup>†</sup> | HR/RR<sup>‡</sup> (95%/96% CI) | p-value | n<sup>i</sup> | Person-time<sup>ii</sup> | Incidence rate<sup>†</sup> | HR/RR<sup>‡</sup> (95%/96% CI) | p-value | n<sup>i</sup> | Person-time<sup>ii</sup> | Incidence rate<sup>†</sup> | HR/RR<sup>‡</sup> (95%/96% CI) | p-value | n<sup>i</sup> | Person-time<sup>ii</sup> | Incidence rate<sup>†</sup> |
| **Primary Outcomes** |  |  |  |  |  |  |  |  |  |  |  |  |  |  |  |  |  |  |
| Nutritional recovery | 144 | 8767 | 599.93 | 1.23 (0.97, 1.57) | 0.083 | 130 | 10106 | 469.84 | 0.94 (0.74, 1.20) | 0.607 | 126 | 10091.9 | 456.02 | 0.91 (0.71, 1.17) | 0.462 | 131 | 9792 | 489.39 |
| Moderate-to-severe diarrhea | 64 | 24545 | 95.17 | 1.26 (0.86, 1.87) | 0.215 | 54 | 23961 | 82.26 | 1.09 (0.73, 1.64) | 0.653 | 63 | 23718.9 | 96.95 | 1.25 (0.84, 1.85) | 0.248 | 50 | 24230 | 75.32 |
| **Secondary Outcomes** |  |  |  |  |  |  |  |  |  |  |  |  |  |  |  |  |  |  |
| Hospitalization or death | 13 | 24350 | 19.50 | 0.70 (0.34, 1.44) | 0.334 | 15 | 23671 | 23.15 | 0.84 (0.42, 1.67) | 0.624 | 17 | 22969.9 | 27.03 | 0.97 (0.50, 1.88) | 0.930 | 18 | 23683 | 27.76 |
| Hospitalization alone | 12 | 24353 | 18.00 | 0.78 (0.36, 1.67) | 0.520 | 11 | 23646 | 16.99 | 0.74 (0.34, 1.61) | 0.448 | 15 | 22969.9 | 23.85 | 1.03 (0.50, 2.11) | 0.936 | 15 | 23660 | 23.16 |
| Severe diarrhea | 10 | 24835 | 14.70 | 1.23 (0.49, 3.23) | 0.661 | 8 | 24223 | 12.05 | 1.01 (0.37, 2.74) | 0.984 | 10 | 23994.9 | 15.21 | 1.02 (0.38, 2.77) | 0.969 | 8 | 24462 | 11.94 |
| Dysentery | 2 | 24907 | 2.93 | 0.33 (0.05, 1.42) | 0.172 | 8 | 24255 | 12.04 | 1.35 (0.47, 4.09) | 0.580 | 7 | 24046.9 | 10.63 | 0.85 (0.24, 2.82) | 0.788 | 6 | 24512 | 8.93 |
| Diarrhea (any severity) | 157 | 24207 | 236.73 | 1.13 (0.90, 1.42) | 0.289 | 158 | 23629 | 244.06 | 1.17 (0.93, 1.47) | 0.186 | 163 | 23353.9 | 254.75 | 1.20 (0.96, 1.51) | 0.111 | 137 | 23911 | 209.13 |
| Medically attended diarrhea over follow-up | 26 | 24787 | 38.29 | 0.85 (0.50, 1.44) | 0.551 | 25 | 24168 | 37.76 | 0.84 (0.49, 1.43) | 0.521 | 27 | 23938.9 | 41.17 | 0.85 (0.50, 1.44) | 0.545 | 30 | 24367 | 44.94 |
| Difference in cumulative duration of diarrhea over follow-up (days)<sup>iii</sup> | - | - | - | 0.18 (-0.90, 1.26) | 0.724 | - | - | - | -0.13 (-1.21, 0.96) | 0.811 | - | - | - | 0.29 (-0.80, 1.38) | 0.610 | - | - | - |

<sup>i</sup>n=number of incident (> 48hours after a diarrhea-free period) moderate-to-severe diarrhea episodes or number of nutritional recovery events (consecutive MUAC measurements $\geq 12.5$ cm); Diarrheal episode defined as $\geq 3$ liquid or semiliquid stools per day after at least 2 diarrhea free days; Moderate to severe diarrhea defined using CODA revised cut-off severity score 3+ as described in SAP
<sup>ii</sup>Person-time in days.
<sup>†</sup> Per 100 child-years
<sup>‡</sup> Hazard Ratio (HR) from cox-proportional hazard model adjusted for inpatient/outpatient status for nutritional recovery outcome and relative risk (RR) for moderate-to-severe diarrhea from Poisson regression model with corresponding 95% CI (nutritional recovery) or 96% CI (moderate-to-severe diarrhea; 96% CI to account for the alpha spent at the interim analysis)

Children with severe wasting randomized to the combination arm had a significantly higher recovery rate than those randomized to the placebo (HR: 2.76, 95% CI 1.49, 5.09; p=0.001; **Figure 3** and **Supp Table 3**) with a median time to recovery of 98 days [IQR: 57.0, 126.0] in the placebo, 84.0 days [IQR: 56.0, 98.5] in the combination **(Supp Table 2)**. Children with severe wasting randomized to lactoferrin alone and lysozyme alone had nutritional recovery rates similar to placebo (HR: 1.29, 95% CI 0.69, 2.41; p=0.427 and HR: 0.80, 95% CI: 0.40, 1.60; p=0.530, respectively). At 16-weeks, 61.8% of children with severe wasting in the placebo, 85.2% in the combination, 58.8% in the lactoferrin, and 48.1% in the lysozyme arms had made a full nutritional recovery. There was no evidence to suggest a difference in nutritional recovery among moderately wasted children, with a high proportion (92.6%-96.8%) of children in all four arms making a full nutritional recovery and a median time to recovery of 42-days across all arms.

**Figure 3:**
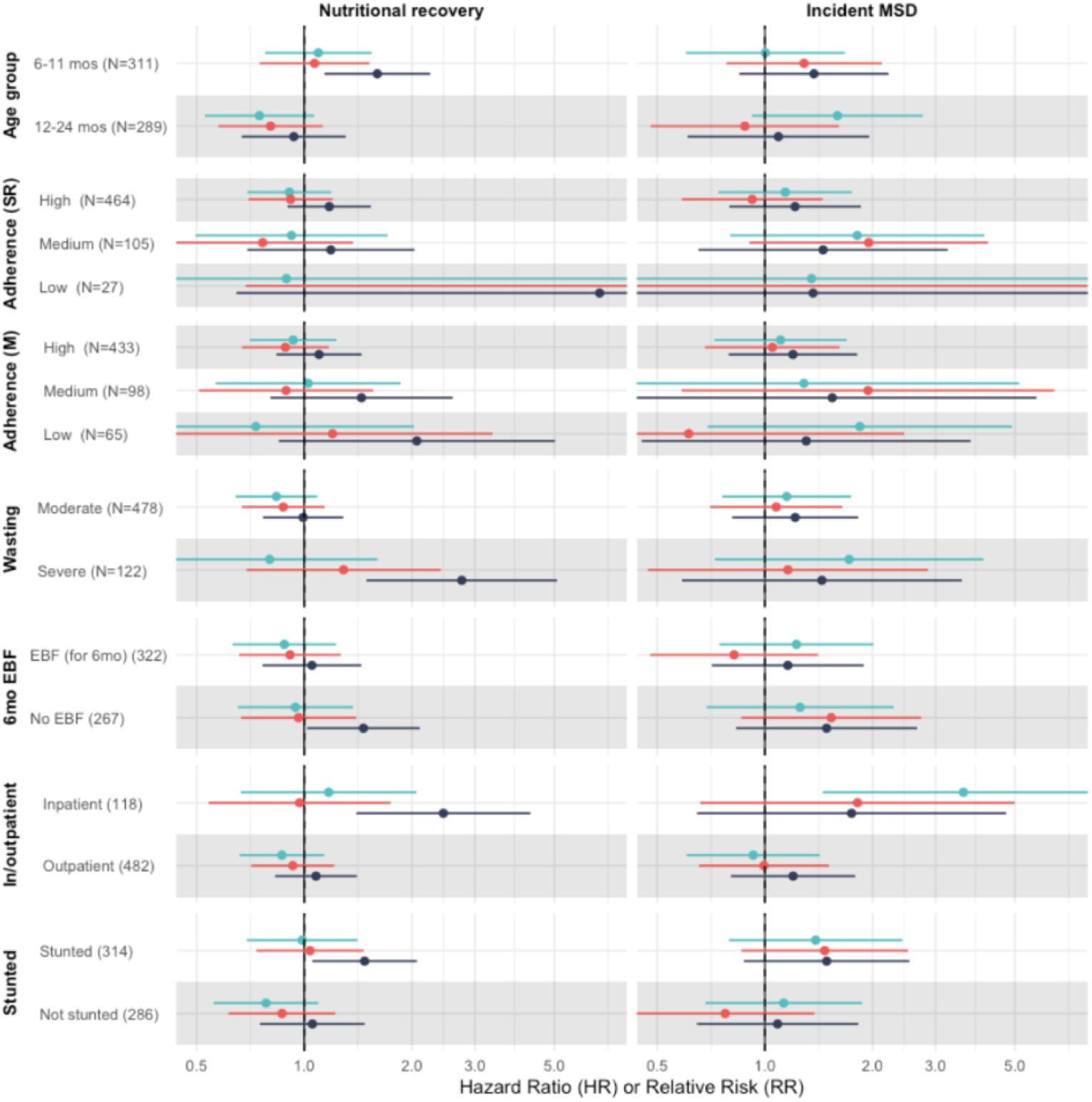
Intervention effectiveness on nutritional recovery and incidence of moderate−to−severe diarrhea across subgroups. RR (95% CI) for incident MSD are the relative risks comparing moderate−to−severe diarrhea (MSD) between each intervention arm and placebo arm obtained from Poisson regression model; HR (95% CI) for the nutritional recovery are the hazard ratio between each intervention arm and placebo arm obtained from Cox Proportional hazard regression model adjusted for outpatient/inpatient status at enrolment; Cox proportional models for in/outpatient status is not adjusted for any covariates. Children who did not have any information on adherence to the intervention are excluded from the subgroup analysis for Self-reported (SR) and Measured (M) adherence.

Subgroup analyses revealed that in addition to children with severe wasting, the combination arm had the greatest impact on nutritional recovery among children who were stunted, children who had been admitted to inpatient wards, younger children (6-11 months vs. 12-24 months), and those not exclusively breastfed for the first 6 months of life (**Figure 3, Supp table 3**). However, there was substantial overlap in all the characteristics among the subgroup of children in whom the combination arm had benefit (**Supp Fig 1**).

### Incidence of MSD

The incidence of MSD during follow-up, the primary diarrhea outcome, was 75.3 episodes per 100 child-years in the placebo arm, and there was no difference in MSD episodes between the intervention arms and placebo (Table 2). There was also no evidence of synergy with lactoferrin and lysozyme on MSD (Wald test for the interaction term p=0.779). The combined impact of lactoferrin, alone or in combination with lysozyme, compared to placebo was 1.05 (96% CI: 0.81, 1.36), while the combined impact of lysozyme was 1.20 (96%CI 0.93, 1.56). Subgroup analyses did not reveal major differences in MSD rates between arms other than a higher incidence of MSD in inpatient children randomized to lysozyme compared to placebo (HR 3.59, 95%CI: 1.45, 8.89; **Figure 3**, **Supp Table 3**).

### Secondary Outcomes

Secondary outcomes, including hospitalization or death, medically attended diarrhea, and other definitions of diarrhea were not meaningfully difference by randomization arm by randomization arm (**Supp Table 4**). In children with severe wasting, there was a modest, albeit not statistically significant reduction in hospitalization between children randomized to the combination arm and the placebo (HR: 0.27 [95%CI: 0.06-1.26, p=0.095) bur no such effect was found for other outcomes (**Supp Figure 2**).

### Per protocol analysis

The per protocol (PP) analysis excluded children who were lost to follow-up (defined as missing all follow-up visits), those who had ≤99% adherence by self-reporting, and those who withdrew consent leaving 399 children (Supp Figure 2). With the reduced sample size, the magnitude of associations remained similar to the ITT, however the combination arm no longer reached marginal statistical significance in terms of improved time to nutritional recovery (HR 1.19 [95%CI: 0.89, 1.58, p=0.235], **Supp Table 4**). Subgroup analyses also did not reveal differences in outcomes between randomization arms stratified by high and low adherence in either the ITT (**Figure 3**) or PP analyses (data not shown).

### Adverse events

There was no evidence of differential severe or mild adverse event rates across the arms. Eleven children (1.8%) died during trial follow-up (3 placebo, 2 combination, 4 lactoferrin, 2 lysozyme) and 8 (72.7%) deaths occurred at home **(Table 3)**. The listed causes of death for the only child with a death certificate was: hypothermia, severe acute malnutrition, and acute kidney injury. No deaths were considered likely related to the IP but one death in the lactoferrin arm was considered possibly related because the child died of comorbid SAM, diarrhea and renal failure during the active course of IP. The safety reviewers noted the relationship between lactoferrin and renal failure was unknown, but also noted the child’s other morbidities to be likely causes of death. There were 54 life threatening events (9%), all of which were hospitalizations. Life threatening events, which were largely hospitalizations for acute illness, were slightly lower in the combination (n=11, 7.2%) and lactoferrin (n=11, 7.4%) arms than the lysozyme (n=16, 10.7%) and placebo (n=16, 10.7%) arm.

**Table 3:** Adverse events by randomization arm.

| Outcome | Overall<br>(N=600) | Lactoferrin +<br>Lysozyme Arm<br>(N=152) | Lactoferrin<br>(N=149) | Lysozyme Arm<br>(N=149) | Placebo Arm<br>(N=150) |
| --- | --- | --- | --- | --- | --- |
| **Serious AE total** | 65 | 13 | 15 | 18 | 19 |
| Grade V: Death | 11 | 2<sup>i</sup> | 4<sup>ii</sup> | 2<sup>iii</sup> | 3<sup>iv</sup> |
| Grade IV: Life threatening | 54 | 11 | 11 | 16 | 16 |
| **AEs total†** | 2608 | 646 | 627 | 691 | 675 |
| Grade III: Severe | 17 | 5 | 5 | 4 | 3 |
| Grade II: Moderate† | 639 | 171 | 156 | 163 | 163 |
| Grade I: Mild† | 1952 | 470 | 466 | 524 | 509 |

i. Causes of deaths: (1) Aspiration pneumonia with tuberculosis and HIV, (2) Unknown
ii. Causes of deaths: (1) Unknown, (2) Acute Kidney Injury, severe anemia, severe acute malnutrition, (3) Unknown, (4) Unknown
iii. Causes of deaths: (1) Pneumonia, (2) Unknown
iv. Causes of deaths: (1) Unknown, (2) Unknown, (3) Unknown
† Percentages reflect the distribution of events within each study arm rather than the proportion of participants affected.

## Discussion

This is the first trial of lactoferrin and lysozyme among wasted children and supplementation with both proteins in combination showed some evidence of a faster recovery from wasting but no evidence of an effect on moderate-to-severe diarrhea incidence. The effect on recovery from wasting was statistically significant among severely wasted children, with a clinically meaningful two-week reduction in the duration of treatment. These children tended to have a higher recovery rate and fewer hospitalizations, suggesting a potential benefit of this combined intervention for this high-risk group. There was no evidence of benefit among children with moderate wasting, a group with high recovery rates across all arms of the trial indicating that they had little need for novel interventions. Our findings suggests that bioactive molecules, like lactoferrin and lysozyme, may not be necessary in the management of moderate wasting but could be useful adjuvants to existing severe wasting treatment.

The management of severe wasting with lipid-based nutritional supplements is expensive, costing up to $300 per-child-treated.^23,24^ However, the large reduction in morbidity and mortality associated with successful wasting management offsets these high costs to make these programs highly cost-effective. Integration of safe and temperature stable bioactive molecules in nutritional supplements may add some cost, but cost savings from a two week reduction in time in the program, as well as reduced hospitalizations which can exceed $200 per inpatient stay^25^, likely exceed any added cost.

Both proteins appeared to be necessary for benefit, perhaps due to synergy between lactoferrin’s ability to sequester iron away from bacterial pathogens and weaken outer cell members for lysozyme to more easily access, and break-down, cell walls^26,27^. That this combination has been shown to reduce hospitalization and reduce enteric measures of inflammation in Malawian children^13^, further substantiates our findings. Prolonged enteric and systemic inflammation in children with severe wasting is implicated in poor recovery and relapse^28,29^ and bioactive molecules targeting inflammation and gut barrier function, such as those tested here and elsewhere^30,31^ hold promise for addressing the enteric dysfunction all too common among wasted children.

Diarrhea incidence rates did not differ across the arms, suggesting that any potential benefits among children with severe wasting cannot be attributed to diarrheal prevention or reduced severity. In Peru, six-months of twice daily lactoferrin given to 12-23 month olds had no effect on the primary outcome of diarrhea incidence ^16^ but had modest improvements in longitudinal prevalence. Smaller trials of lactoferrin and lysozyme combination among 5-33 month Peruvian children, ^17^ and lactoferrin alone in Chinese children aged 4-6 months,^15^ showed similarly inconsistent diarrhea benefits. Incidence of MSD in our trial population was nearly 100 per 100 child-years (and over 200 episodes of any severity diarrhea per 100-child years). The high burden of diarrheal episodes among children with wasting both during recovery and post discharge from malnutrition programs is well characterized, as is diarrhea’s association with relapse. ^32,33^. It is possible that asymptomatic enteric infections were reduced by the combination of lactoferrin and lysozyme and that this effect mediated the association between the combination arm and nutritional recovery without impacting diarrhea. Asymptomatic enteric infections have been shown to increase. The cumulative enteric infection burden in early life is more predictive of linear growth faltering, a precursor to stunting, than cumulative diarrheal episodes. These pathogens increase enteric inflammation^14^ and subsequent malabsorption of nutrients and systemic inflammation.

This was rigorously conducted, blinded, randomized control trial with excellent adherence and a high follow-up rate. However we were limited by several factors. The sample size for this study was relatively small, therefore findings among subpopulations, even those pre-specified such as children with severe wasting, should temper causal inference. Similarly, secondary outcomes such as hospitalization, death, and medically attended diarrhea were relatively rare and as secondary to the primary outcomes, should be interpreted with caution. While we assessed adherence in two, independent ways, both self-report and sachet collection are imperfect and may have led to inflated adherence estimates.

## Conclusion

This trial suggests that lactoferrin and lysozyme in combination may shorten the duration of treatment for children with severe wasting and diarrhea, but they did not benefit moderately wasted children. These data suggest interventions targeting the enteric system may benefit severely wasted children with diarrhea, but confirmatory studies validating this effect are needed prior to implementation and policy change. Diarrhea remains a common condition experienced by wasted children and interventions preventing such complications remain needed.

## Data Availability

All data produced in the present study are available upon reasonable request to the authors

## Acknowledgments

The authors thank the children who participated in these studies and their families, and the dedicated physicians, nurses, scientists, and staff at each hospital for their dedication and outstanding performance of clinical and laboratory study activities. This study was funded by the National Institute of Health R01HD103642. We are also grateful to the members of our data safety and monitoring board: Dr. Helen Powell-University of Maryland School of Medicine; Dr. Jeffrey Pernica, McMaster University; Dr. Ayesa De Costa, World Health Organization; Dr. Phelgona Otieno, Kenya Medical Research Institute; Mary Amondi, International AIDS Vaccine Initiative (IAVI). We thank FrieslandCampina for their donation of lactoferrin for the trial.

## Appendix

**Supplementary Table 1:** Participant reported and objectively measured adherence to the investigation product (IP) over 16 weeks.

|  | **Overall**<br>**(n=596)<sup>§</sup>** | **Lactoferrin+**<br>**Lysozyme**<br>**(n=152)** | **Lactoferrin**<br>**(n=148)** | **Lysozyme**<br>**(n=146)** | **Placebo (n=150)** |
| --- | --- | --- | --- | --- | --- |
| **Self-reported Adherence<sup>†</sup>** |  |  |  |  |  |
| Duration of IP consumption (days)<sup>††</sup> | 112 [111, 112] | 112 [111, 112] | 112 [111, 112] | 112 [112, 112] | 112 [110, 112] |
| High (n, %) | 464 (77.9%) | 118 (77.6%) | 117 (79.1%) | 118 (80.8%) | 111 (74.0%) |
| Medium (n, %) | 105 (17.6%) | 27 (17.8%) | 25 (16.9%) | 20 (13.7%) | 33 (22.0%) |
| Low (n, %) | 27 (4.5%) | 7 (4.6%) | 6 (4.1%) | 8 (5.5%) | 6 (4.0%) |
| **Measured Adherence<sup>‡</sup>** |  |  |  |  |  |
| Duration of IP consumption (days)<sup>††</sup> | 112 [112, 113] | 112 [112, 112] | 112 [112, 113] | 112 [112, 113] | 112 [112, 113] |
| High (n, %) | 433 (72.7%) | 113 (74.3%) | 106 (71.6%) | 108 (74.0%) | 106 (70.7%) |
| Medium (n, %) | 98 (16.4%) | 20 (13.2%) | 29 (19.6%) | 20 (13.7%) | 28 (18.7%) |
| Low (n, %) | 65 (10.9%) | 17 (11.2%) | 14 (9.5%) | 17 (11.6%) | 17 (11.3%) |

<sup>§</sup>There were three deaths and one withdrawal before the Week 2 visit when adherence data was collected; these four children have missing data for adherence. If a child did not attend any given follow up visits during the 16-week course of IP, it was assumed that the child did not consume the IP for that visit.
<sup>††</sup>Values are Median [IQR]
<sup>†</sup>Self-reported adherence (16-week overall adherence) = Number of IP sachets consumed in 112 days divided by 112.
<sup>‡</sup>Measured adherence (16-week overall adherence) = Number of visits for which the caregiver returned at least 14 IP sachets at each visit divided by eight, the total number of scheduled visits the child was expected to attend during the course of the study.
High adherence defined as >99%, medium as 85-99%, and low as <85%.

**Supplementary Table 2:** Proportion of children who recovered from wasting by randomization arm and wasting severity at enrollment.

|  | Total (N=600) | Lactoferrin +<br>Lysozyme Arm<br>(N =152) | Lactoferrin Arm<br>(N =149) | Lysozyme Arm<br>(N =149) | Placebo Arm<br>(N =150) |
| --- | --- | --- | --- | --- | --- |
| Recovered, N (%) at any point during follow-up<sup>†, i</sup> |  |  |  |  |  |
| **Overall** | 531 (88.5%) | 144 (94.7%) | 130 (87.2%) | 126 (84.6%) | 131 (87.3%) |
| Moderate wasting<sup>‡</sup> | 454 (95.0%) | 121 (96.8%) | 110 (95.7%) | 113 (92.6%) | 110 (94.8%) |
| Severe wasting | 77 (63.1%) | 23 (85.2%) | 20 (58.8%) | 13 (48.1%) | 21 (61.8%) |
| Days to recovery among those who recovered, Median (IQR) |  |  |  |  |  |
| **Overall** | 42.0 [29.5, 70.0] | 42.0 [29.0, 60.8] | 42.0 [42.0, 70.0] | 42.0 [31.3, 70.0] | 42.0 [28.0, 70.0] |
| Moderate wasting | 42.0 [28.0, 56.0] | 42.0 [28.0, 56.0] | 42.0 [42.0, 56.8] | 42.0 [28.0, 56.0] | 42.0 [28.0, 56.0] |
| Severe wasting | 84.0 [56.0, 113] | 84.0 [56.0, 98.5] | 84.0 [76.8, 102] | 98.0 [56.0, 112] | 98.0 [57.0, 126] |

Severe wasting: MUAC of $\leq 11.5$ cm; moderate wasting: MUAC of 11.5-12.4cm.
<sup>†</sup>Wasting recovery defined as the number of days since enrollment to the date of the second of two consecutive fortnightly MUAC measurements $\geq 12.5$ cm. Number of children who relapsed after nutritional recovery during the study= 18 (3.0%).
<sup>i</sup>For participants who did not reach nutritional recovery, 11 (15.9%) were due to death, 2 (2.9%) were due to withdrawal, 3 (4.3%) were due to being untraceable, 2 (2.9%) were due to other reasons for termination (e.g, non-compliance), and 51 (73.9%) completed the study prior to reaching recovery.
<sup>‡</sup>Number of children who were enrolled as moderately wasted and became severe during study (at exit) = 1 (0.2%)

**Supplementary Table 3:**
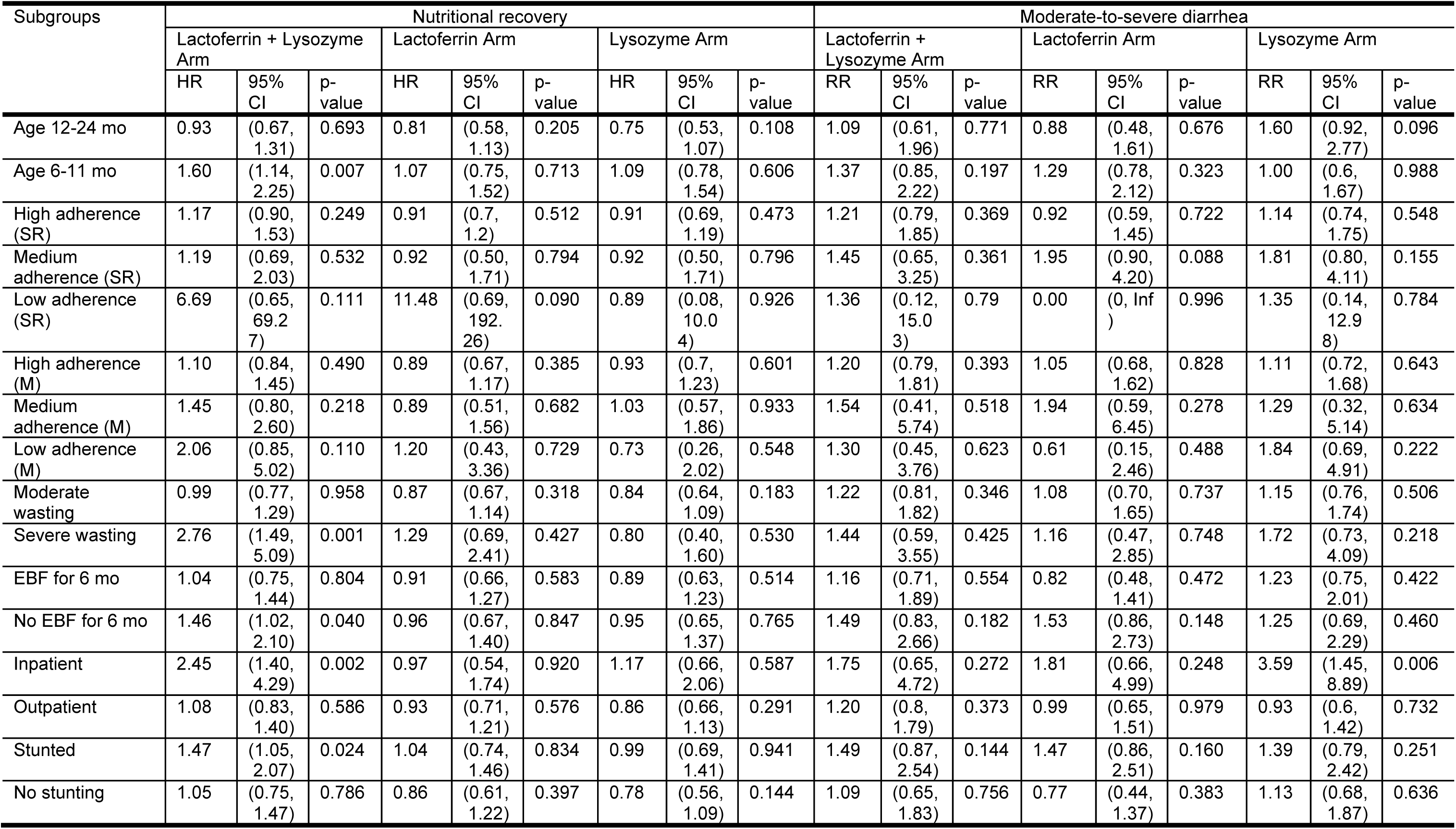
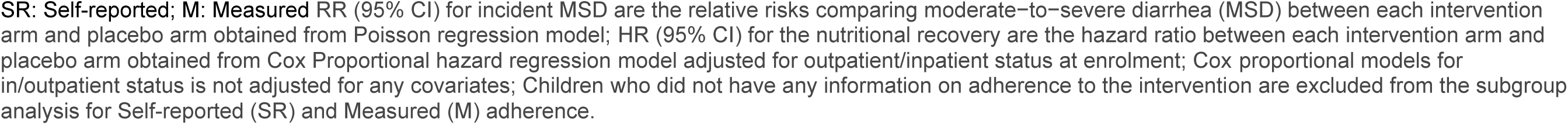
Intervention effectiveness within subgroups on nutritional recovery and incidence of moderate−to−severe diarrhea.

| Subgroups | Nutritional recovery |  |  |  |  |  |  |  |  | Moderate-to-severe diarrhea |  |  |  |  |  |  |  |  |
| --- | --- | --- | --- | --- | --- | --- | --- | --- | --- | --- | --- | --- | --- | --- | --- | --- | --- | --- |
|  | Lactoferrin + Lysozyme Arm |  |  | Lactoferrin Arm |  |  | Lysozyme Arm |  |  | Lactoferrin + Lysozyme Arm |  |  | Lactoferrin Arm |  |  | Lysozyme Arm |  |  |
|  | HR | 95% CI | p-value | HR | 95% CI | p-value | HR | 95% CI | p-value | RR | 95% CI | p-value | RR | 95% CI | p-value | RR | 95% CI | p-value |
| Age 12-24 mo | 0.93 | (0.67, 1.31) | 0.693 | 0.81 | (0.58, 1.13) | 0.205 | 0.75 | (0.53, 1.07) | 0.108 | 1.09 | (0.61, 1.96) | 0.771 | 0.88 | (0.48, 1.61) | 0.676 | 1.60 | (0.92, 2.77) | 0.096 |
| Age 6-11 mo | 1.60 | (1.14, 2.25) | 0.007 | 1.07 | (0.75, 1.52) | 0.713 | 1.09 | (0.78, 1.54) | 0.606 | 1.37 | (0.85, 2.22) | 0.197 | 1.29 | (0.78, 2.12) | 0.323 | 1.00 | (0.6, 1.67) | 0.988 |
| High adherence (SR) | 1.17 | (0.90, 1.53) | 0.249 | 0.91 | (0.7, 1.2) | 0.512 | 0.91 | (0.69, 1.19) | 0.473 | 1.21 | (0.79, 1.85) | 0.369 | 0.92 | (0.59, 1.45) | 0.722 | 1.14 | (0.74, 1.75) | 0.548 |
| Medium adherence (SR) | 1.19 | (0.69, 2.03) | 0.532 | 0.92 | (0.50, 1.71) | 0.794 | 0.92 | (0.50, 1.71) | 0.796 | 1.45 | (0.65, 3.25) | 0.361 | 1.95 | (0.90, 4.20) | 0.088 | 1.81 | (0.80, 4.11) | 0.155 |
| Low adherence (SR) | 6.69 | (0.65, 69.27) | 0.111 | 11.48 | (0.69, 192.26) | 0.090 | 0.89 | (0.08, 10.04) | 0.926 | 1.36 | (0.12, 15.03) | 0.79 | 0.00 | (0, Inf) | 0.996 | 1.35 | (0.14, 12.98) | 0.784 |
| High adherence (M) | 1.10 | (0.84, 1.45) | 0.490 | 0.89 | (0.67, 1.17) | 0.385 | 0.93 | (0.7, 1.23) | 0.601 | 1.20 | (0.79, 1.81) | 0.393 | 1.05 | (0.68, 1.62) | 0.828 | 1.11 | (0.72, 1.68) | 0.643 |
| Medium adherence (M) | 1.45 | (0.80, 2.60) | 0.218 | 0.89 | (0.51, 1.56) | 0.682 | 1.03 | (0.57, 1.86) | 0.933 | 1.54 | (0.41, 5.74) | 0.518 | 1.94 | (0.59, 6.45) | 0.278 | 1.29 | (0.32, 5.14) | 0.634 |
| Low adherence (M) | 2.06 | (0.85, 5.02) | 0.110 | 1.20 | (0.43, 3.36) | 0.729 | 0.73 | (0.26, 2.02) | 0.548 | 1.30 | (0.45, 3.76) | 0.623 | 0.61 | (0.15, 2.46) | 0.488 | 1.84 | (0.69, 4.91) | 0.222 |
| Moderate wasting | 0.99 | (0.77, 1.29) | 0.958 | 0.87 | (0.67, 1.14) | 0.318 | 0.84 | (0.64, 1.09) | 0.183 | 1.22 | (0.81, 1.82) | 0.346 | 1.08 | (0.70, 1.65) | 0.737 | 1.15 | (0.76, 1.74) | 0.506 |
| Severe wasting | 2.76 | (1.49, 5.09) | 0.001 | 1.29 | (0.69, 2.41) | 0.427 | 0.80 | (0.40, 1.60) | 0.530 | 1.44 | (0.59, 3.55) | 0.425 | 1.16 | (0.47, 2.85) | 0.748 | 1.72 | (0.73, 4.09) | 0.218 |
| EBF for 6 mo | 1.04 | (0.75, 1.44) | 0.804 | 0.91 | (0.66, 1.27) | 0.583 | 0.89 | (0.63, 1.23) | 0.514 | 1.16 | (0.71, 1.89) | 0.554 | 0.82 | (0.48, 1.41) | 0.472 | 1.23 | (0.75, 2.01) | 0.422 |
| No EBF for 6 mo | 1.46 | (1.02, 2.10) | 0.040 | 0.96 | (0.67, 1.40) | 0.847 | 0.95 | (0.65, 1.37) | 0.765 | 1.49 | (0.83, 2.66) | 0.182 | 1.53 | (0.86, 2.73) | 0.148 | 1.25 | (0.69, 2.29) | 0.460 |
| Inpatient | 2.45 | (1.40, 4.29) | 0.002 | 0.97 | (0.54, 1.74) | 0.920 | 1.17 | (0.66, 2.06) | 0.587 | 1.75 | (0.65, 4.72) | 0.272 | 1.81 | (0.66, 4.99) | 0.248 | 3.59 | (1.45, 8.89) | 0.006 |
| Outpatient | 1.08 | (0.83, 1.40) | 0.586 | 0.93 | (0.71, 1.21) | 0.576 | 0.86 | (0.66, 1.13) | 0.291 | 1.20 | (0.8, 1.79) | 0.373 | 0.99 | (0.65, 1.51) | 0.979 | 0.93 | (0.6, 1.42) | 0.732 |
| Stunted | 1.47 | (1.05, 2.07) | 0.024 | 1.04 | (0.74, 1.46) | 0.834 | 0.99 | (0.69, 1.41) | 0.941 | 1.49 | (0.87, 2.54) | 0.144 | 1.47 | (0.86, 2.51) | 0.160 | 1.39 | (0.79, 2.42) | 0.251 |
| No stunting | 1.05 | (0.75, 1.47) | 0.786 | 0.86 | (0.61, 1.22) | 0.397 | 0.78 | (0.56, 1.09) | 0.144 | 1.09 | (0.65, 1.83) | 0.756 | 0.77 | (0.44, 1.37) | 0.383 | 1.13 | (0.68, 1.87) | 0.636 |

SR: Self-reported; M: Measured RR (95% CI) for incident MSD are the relative risks comparing moderate-to-severe diarrhea (MSD) between each intervention arm and placebo arm obtained from Poisson regression model; HR (95% CI) for the nutritional recovery are the hazard ratio between each intervention arm and placebo arm obtained from Cox Proportional hazard regression model adjusted for outpatient/inpatient status at enrolment; Cox proportional models for in/outpatient status is not adjusted for any covariates; Children who did not have any information on adherence to the intervention are excluded from the subgroup analysis for Self-reported (SR) and Measured (M) adherence.

**Supplementary Figure 1:**
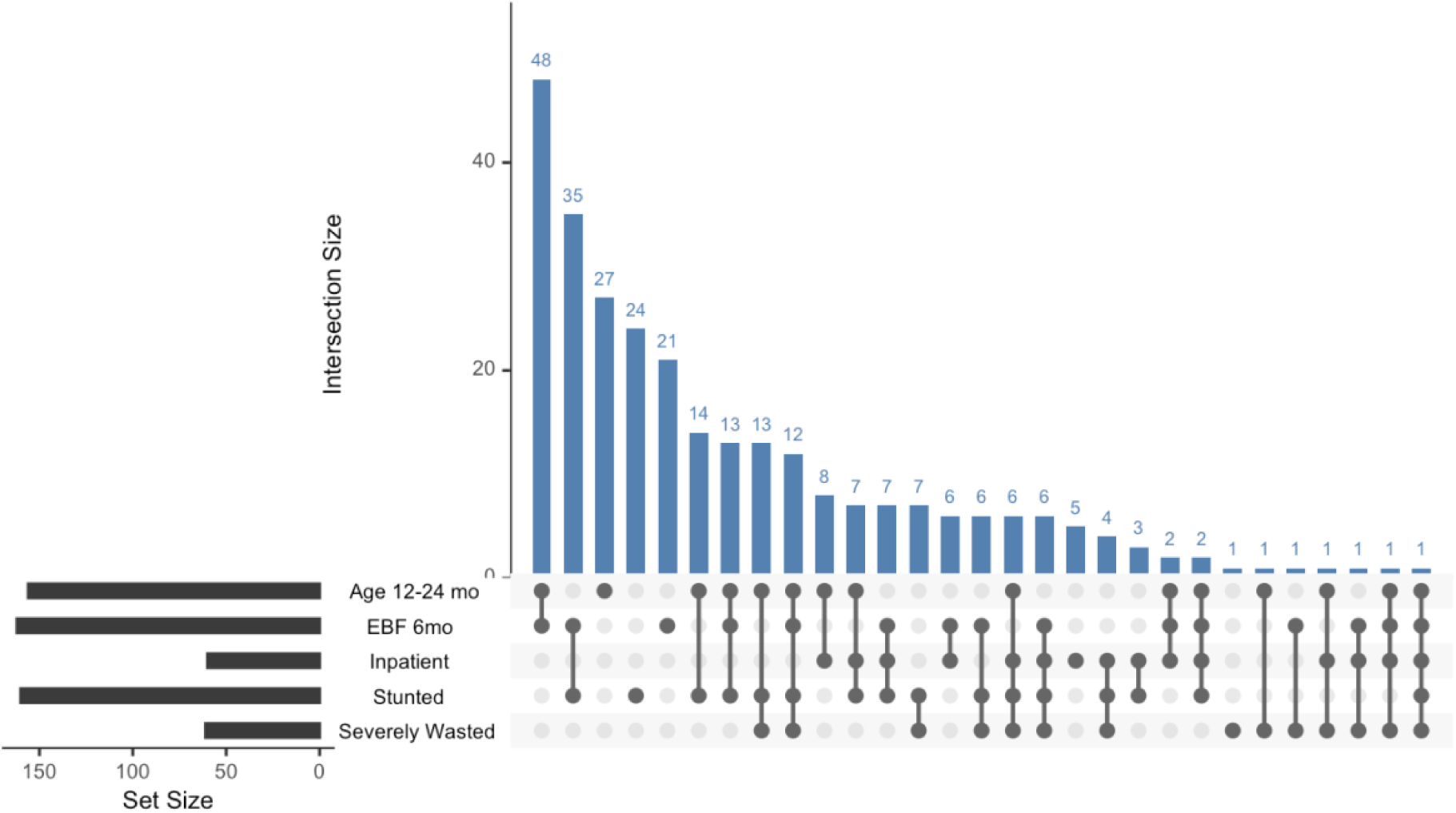
Distribution of participants included in the lactoferrin + lysozyme and placebo arms by subgroups. UpSet plot of overlapping baseline characteristics among children in the combination and placebo arm, since the combination arm showed a significant impact on nutritional recovery in several subgroup analyses compared to placebo. The horizontal bars on the left show the number of participants in each subgroup: age 12 to 24 months, exclusive breastfeeding for 6 months, inpatient enrollment, stunting, and severe wasting. The filled, connected dots identify each subgroup combination, and the vertical bars represent the number of participants in each corresponding intersection. Only combinations present in the data are shown; children with none of these characteristics are not included in the intersection panel.

**Supplementary Figure 2:**
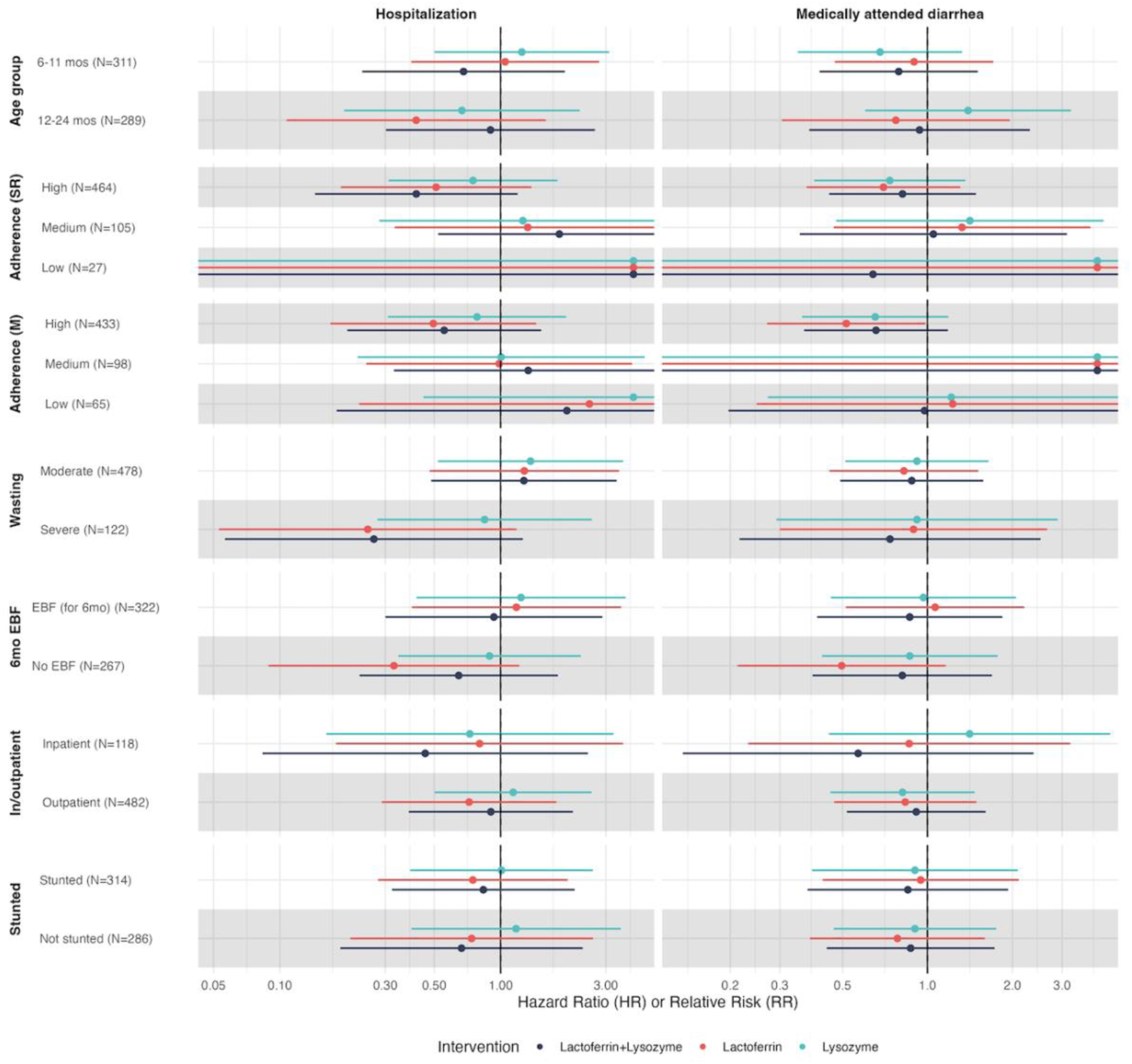
Subgroup analyses among select secondary outcomes.

**Supplementary Figure 2:**
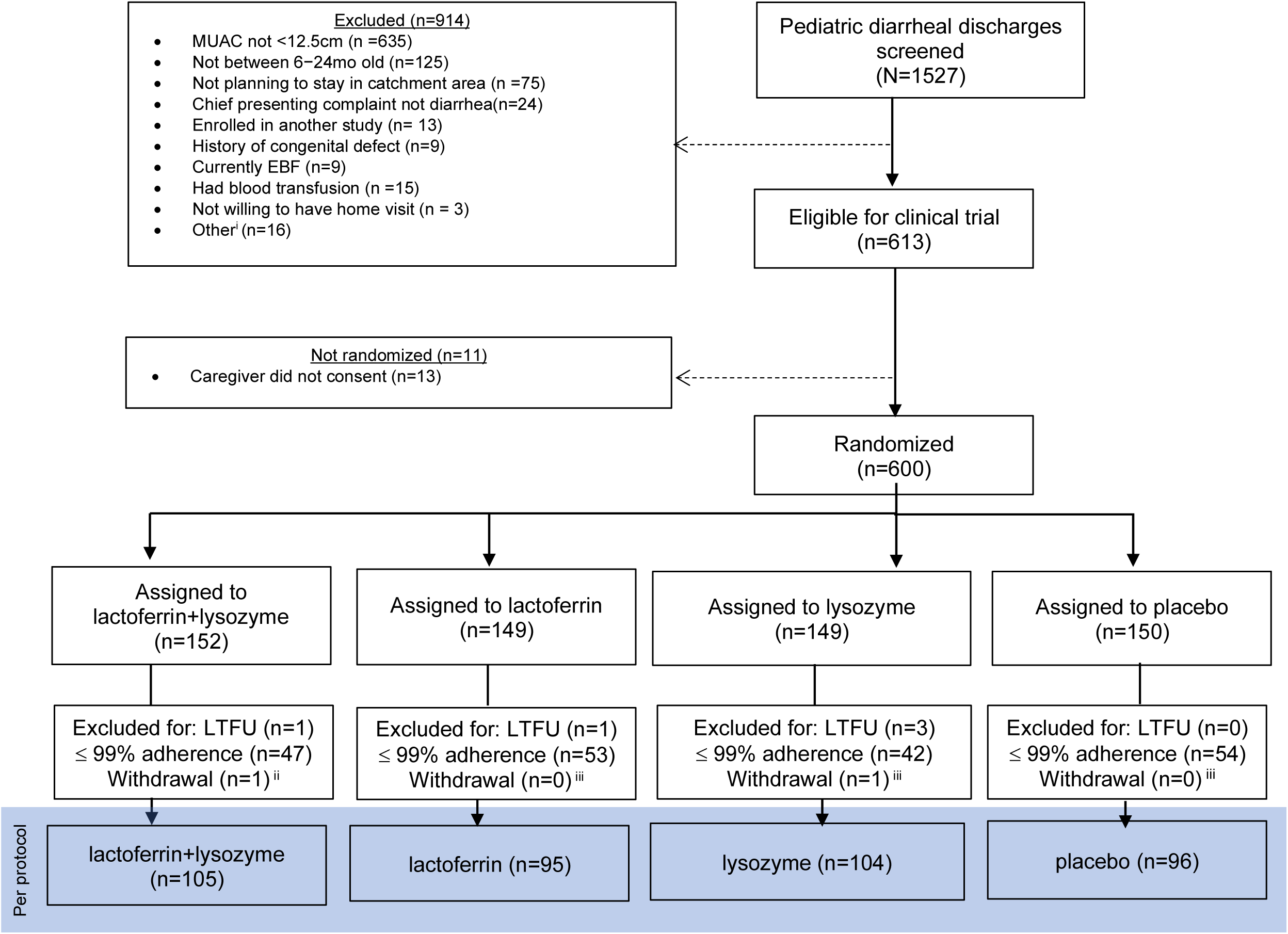
Flowchart of included subjects (per-protocol analysis) • ^i^Includes the following exclusions: Child not deemed clinically stable (n =1); Discharged against medical advice (n = 2); Not willing to prepare study product (n=2); Already enrolled in this study (n= 1); Not willing to provide consent (n =2); Not willing to bring child to clinic (n =2); child was already discharged for at least 2 days when screened (n=2); caregiver did not have time to sit through the whole process of consenting (n=1), caregiver was committed elsewhere (n=2), and caregiver requested to consult with partner who declined enrolment (n=1). • ^ii^The exclusion categories overlap

**Supplementary Table 4:**
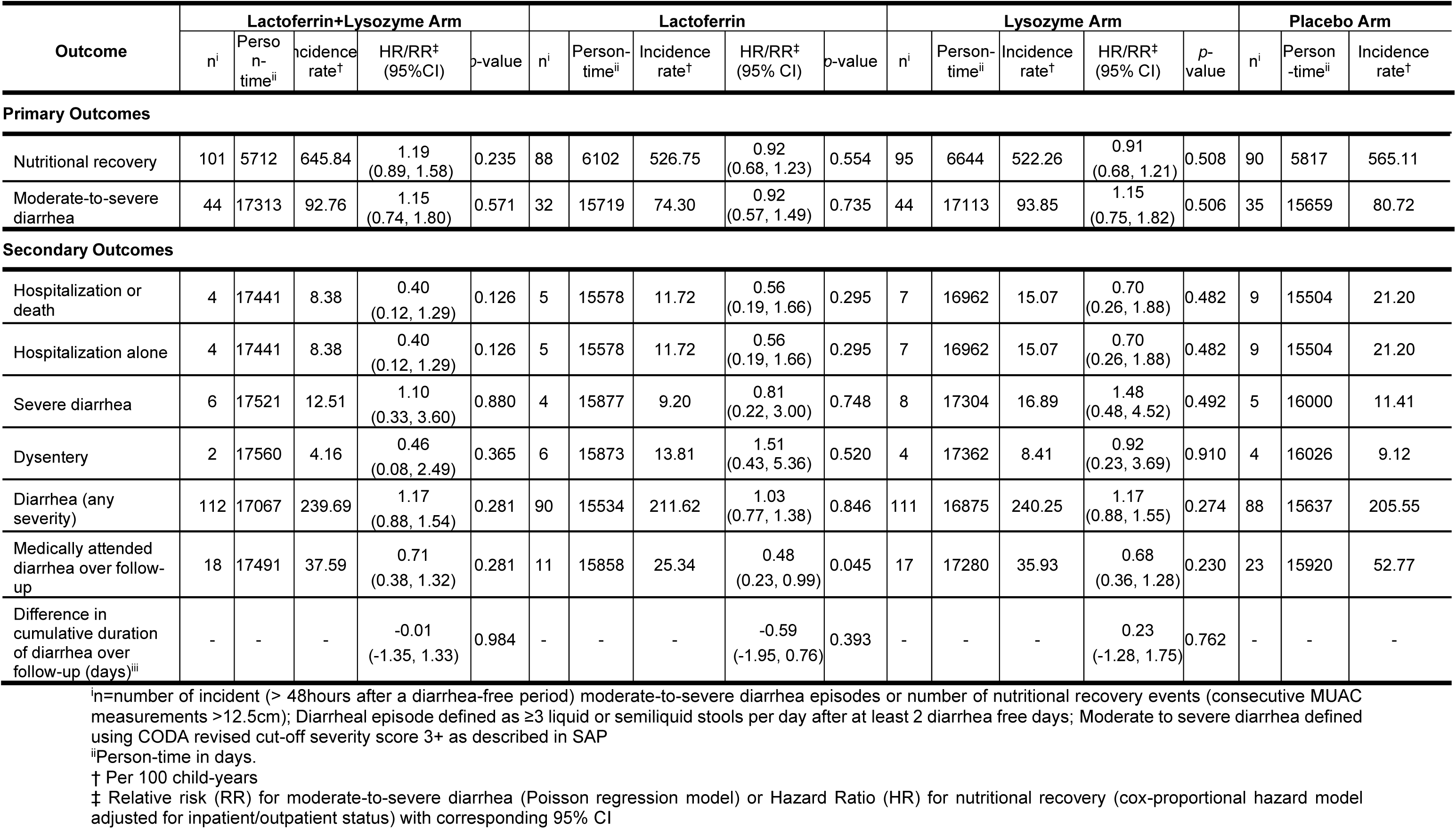
Nutritional recovery and moderate-to-severe diarrhea by randomization arm (per protocol analysis)

| Outcome | Lactoferrin+Lysozyme Arm |  |  |  |  | Lactoferrin |  |  |  |  | Lysozyme Arm |  |  |  |  | Placebo Arm |  |  |
| --- | --- | --- | --- | --- | --- | --- | --- | --- | --- | --- | --- | --- | --- | --- | --- | --- | --- | --- |
|  | n<sup>i</sup> | Person-time<sup>ii</sup> | Incidence rate<sup>†</sup> | HR/RR<sup>‡</sup> (95%CI) | p-value | n<sup>i</sup> | Person-time<sup>ii</sup> | Incidence rate<sup>†</sup> | HR/RR<sup>‡</sup> (95% CI) | p-value | n<sup>i</sup> | Person-time<sup>ii</sup> | Incidence rate<sup>†</sup> | HR/RR<sup>‡</sup> (95% CI) | p-value | n<sup>i</sup> | Person-time<sup>ii</sup> | Incidence rate<sup>†</sup> |
| **Primary Outcomes** |  |  |  |  |  |  |  |  |  |  |  |  |  |  |  |  |  |  |
| Nutritional recovery | 101 | 5712 | 645.84 | 1.19<br>(0.89, 1.58) | 0.235 | 88 | 6102 | 526.75 | 0.92<br>(0.68, 1.23) | 0.554 | 95 | 6644 | 522.26 | 0.91<br>(0.68, 1.21) | 0.508 | 90 | 5817 | 565.11 |
| Moderate-to-severe diarrhea | 44 | 17313 | 92.76 | 1.15<br>(0.74, 1.80) | 0.571 | 32 | 15719 | 74.30 | 0.92<br>(0.57, 1.49) | 0.735 | 44 | 17113 | 93.85 | 1.15<br>(0.75, 1.82) | 0.506 | 35 | 15659 | 80.72 |
| **Secondary Outcomes** |  |  |  |  |  |  |  |  |  |  |  |  |  |  |  |  |  |  |
| Hospitalization or death | 4 | 17441 | 8.38 | 0.40<br>(0.12, 1.29) | 0.126 | 5 | 15578 | 11.72 | 0.56<br>(0.19, 1.66) | 0.295 | 7 | 16962 | 15.07 | 0.70<br>(0.26, 1.88) | 0.482 | 9 | 15504 | 21.20 |
| Hospitalization alone | 4 | 17441 | 8.38 | 0.40<br>(0.12, 1.29) | 0.126 | 5 | 15578 | 11.72 | 0.56<br>(0.19, 1.66) | 0.295 | 7 | 16962 | 15.07 | 0.70<br>(0.26, 1.88) | 0.482 | 9 | 15504 | 21.20 |
| Severe diarrhea | 6 | 17521 | 12.51 | 1.10<br>(0.33, 3.60) | 0.880 | 4 | 15877 | 9.20 | 0.81<br>(0.22, 3.00) | 0.748 | 8 | 17304 | 16.89 | 1.48<br>(0.48, 4.52) | 0.492 | 5 | 16000 | 11.41 |
| Dysentery | 2 | 17560 | 4.16 | 0.46<br>(0.08, 2.49) | 0.365 | 6 | 15873 | 13.81 | 1.51<br>(0.43, 5.36) | 0.520 | 4 | 17362 | 8.41 | 0.92<br>(0.23, 3.69) | 0.910 | 4 | 16026 | 9.12 |
| Diarrhea (any severity) | 112 | 17067 | 239.69 | 1.17<br>(0.88, 1.54) | 0.281 | 90 | 15534 | 211.62 | 1.03<br>(0.77, 1.38) | 0.846 | 111 | 16875 | 240.25 | 1.17<br>(0.88, 1.55) | 0.274 | 88 | 15637 | 205.55 |
| Medically attended diarrhea over follow-up | 18 | 17491 | 37.59 | 0.71<br>(0.38, 1.32) | 0.281 | 11 | 15858 | 25.34 | 0.48<br>(0.23, 0.99) | 0.045 | 17 | 17280 | 35.93 | 0.68<br>(0.36, 1.28) | 0.230 | 23 | 15920 | 52.77 |
| Difference in cumulative duration of diarrhea over follow-up (days)<sup>iii</sup> | - | - | - | -0.01<br>(-1.35, 1.33) | 0.984 | - | - | - | -0.59<br>(-1.95, 0.76) | 0.393 | - | - | - | 0.23<br>(-1.28, 1.75) | 0.762 | - | - | - |

<sup>i</sup>n=number of incident (> 48hours after a diarrhea-free period) moderate-to-severe diarrhea episodes or number of nutritional recovery events (consecutive MUAC measurements >12.5cm); Diarrheal episode defined as ≥3 liquid or semiliquid stools per day after at least 2 diarrhea free days; Moderate to severe diarrhea defined using CODA revised cut-off severity score 3+ as described in SAP
<sup>ii</sup>Person-time in days.
<sup>†</sup> Per 100 child-years
<sup>‡</sup> Relative risk (RR) for moderate-to-severe diarrhea (Poisson regression model) or Hazard Ratio (HR) for nutritional recovery (cox-proportional hazard model adjusted for inpatient/outpatient status) with corresponding 95% CI

## Notes

### Competing Interest Statement

The authors have declared no competing interest.

### Clinical Trial

NCT05519254

### Clinical Protocols

https://bmjopen.bmj.com/content/bmjopen/14/8/e079448.full.pdf

### Author Declarations

The clinical trial which was approved by the University of Washington Institutional Review Board STUDY00011759 and Kenya Medical Research Institutes Scientific and Ethical Review Unit SERU 4274

### Summary of Updates

Corrected Caregiver HIV status in Table 1 -- HIV infection and HIV uninfected numbers were reversed.

